# Making the Match or Breaking it? Values, Perceptions, and Obstacles of Trainees Applying into Physician-Scientist Training Programs

**DOI:** 10.1101/2022.08.27.22278743

**Authors:** M.E. Pepin, Y. Kamal, B.J. Reisman, M.E. Rockman, J.P. Waller

## Abstract

**Background:** Replenishing the physician-scientist workforce remains a central mission of medical education, but the hemorrhaging of qualified trainees threatens the physician-scientist role. Among the barriers facing physician-scientists is the game-like model of residency matching, which applies several flawed assumptions regarding the comparability of applicant qualifications, cohort size, and the institutional breadth of applicants’ training needs.

**Methods:** The current report summarizes the collective views and experiences of physician-scientist trainees following the 2021-2022 application cycle of physician-scientist training programs (PSTPs). We obtained survey-based feedback by 27 PSTP applicants from 17 U.S. medical universities, among whom 85% matched into a PSTP.

**Results:** 93% PSTP applicants recognized “scientific community” as the most important feature of postgraduate training, whereas gender-specific worth was assigned to institutional aspects of personal and/or family support. 65% of respondents found “waiting for interviews” as the most stressful aspect of the application cycle. Half of the survey respondents perceived at least one NRMP policy violation by a PSTP, most of which occurred during post-interview communication. 93% of respondents were contacted by a PSTP following interviews, and one-third admitted to feeling pressured into sharing their ranking preferences.

**Conclusion:** Overall, we believe the values and needs of physician-scientist trainees are poorly represented by the current PSTP selection framework, including inconsistent timelines and communication throughout the process. We propose a series of modifications that, if implemented, would better equip applicants to gauge programs according to the clinical, scientific, and academic communities that we seek to join as academic physician-scientists.

## INTRODUCTION

Physician-scientists represent a dwindling entity within the U.S. biomedical workforce, the purpose of which has long been to lead the transformation of biomedical discoveries into clinical practice. Although NIH-funded predoctoral MD-PhD programs continue to graduate over 600 trainees each year, the nationwide prevalence of tenured physician-scientists continues to decline.^1^ Institutions have therefore begun implementing specialized postgraduate tracks to promote retention of physician-scientist trainees, using research-integrated residency programs, or physician-scientist training programs (PSTPs), to interweave research mentoring and experiences alongside residency training.^2^ Although no studies have yet empirically shown their worth as retention tools, PSTPs offer many aspects of training support that are likely to benefit the long-term success of trainees, including structured mentorship and protected research time.

Despite their advantages, only a small proportion of MD-PhD graduates enter research-oriented residencies, owing in-part to the incapacity of these small programs to accommodate the steady influx of residency-seeking physician-scientists. This bottleneck creates an unnecessary bifurcation in the physician-scientist training “pipeline,” leaving many highly-qualified MD-PhD graduates unsupported during their residency training.^3,4^ The issue has been more recently compounded by the COVID-19 pandemic, which disproportionately burdened female trainees and other underrepresented groups.^5,6^ Strategies are therefore needed to both widen entry into PSTPs and identify the priorities of applicants during this pivotal transition in their professional advancement.

To understand the qualities, values, and struggles facing PSTP-seeking residency applicants, we distributed a post-match survey to identify PSTP applicants’ perceptions of the application, interview, and match processes into – or away from – programs across the United States. Responses highlight that ambiguous and non-standardized communication promotes applicant anxiety and leads to numerous violations in National Residency Matching Program (NRMP) policy.

## METHODS

We distributed a voluntary and anonymous survey to capture the experiences among physician-scientist trainees seeking to enter research-oriented Internal Medicine residency programs, or physician-scientist training programs (PSTPs). This post-match survey was distributed to applicants of at least 1 PSTP during the 2021-2022 cycle. In its preamble, the designation of “PSTP” was explicitly defined as residency tracks that “integrate clinical training in internal medicine residency and fellowship activities over a six to seven-year period of training time,” as described by the American Association of Medical Colleges.^7^

Ethical approval for this work was provided by the Office of the Institutional Review Board at the University of Alabama at Birmingham (IRB-300001128). Participation in the research study was strictly voluntary, participants were allowed to withdraw at any time without repercussion or response tracking. The process involved filling an online survey that took approximately 10 minutes to finish. All responses were kept confidential, and no personally identifying information (name, email address or IP address) was collected. Regardless, all data were stored in a password-protected electronic format and – owing to the small applicant pool and potential for inadvertent identification – results are reported as aggregate values only.

Statistical analyses and data visualizations were done using GraphPad Prism version 9.0 for Macintosh (GraphPad Software, San Diego, CA) and R software, version 4.1.2 (Vienna, Austria). Statistical significance was assigned when *P* < 0.05. All data are represented as mean +/- standard deviation unless otherwise indicated. Statistical significance of pair-wise comparisons was determined using unpaired Student’s *t-*tests following Shapiro-Wilk test for normality. For multiple comparisons testing, correlation analysis via Pearson’s chi-squared analysis was performed in combination with linear multiple linear regression, as follows:

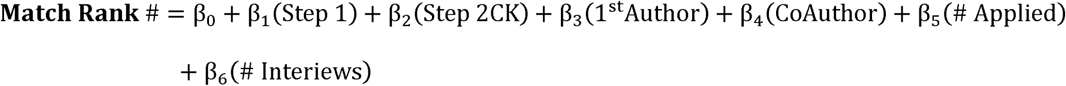

## RESULTS

### Characteristics of applicant survey respondents

To develop a collective sense of applicants’ qualities, values, and experiences during the 2021-2022 application cycle to post-graduate PSTPs, a voluntary survey was distributed in the week following their matching into these programs. The first series of questions collected demographic features, geographical affiliations, and scholastic performances of respondents (**Fig. 1**). From these, 17 undergraduate medical institutions were represented across 17 states in the Southeast, Northeast, and Mid-West (**Fig. 1A**). Most (93%) respondents were graduating MD-PhD trainees, 67% identified as white, and 78% as male (**Fig. 1B**). PSTP applicants scored higher in both Step 1 (Δ = 9.6 pts, *P* = 0.02) and Step 2 CK (Δ = 10.2 pts, *P* = 0.01) examinations relative to the national average, with a 10.7-point increase (*P* = 0.0043) occurring between their Step 1 (240 ± 6) and Step 2 CK (250.7 ± 4.3) scores (**Fig. 1C**). Contrary to national trends, however, the distribution of USMLE Step 1 scores equally represented all quartiles of score performance (**Fig. 1D**). PSTP applicants demonstrated above-average research productivity relative to national averages for categorical internal medicine applicants, entering the residency application with a median of 3 first-authored and 10 co-authored publications (**Fig. 1E**). Additionally, PSTP respondents applied to 20 programs, far fewer than categorical internal medicine applicants entering the same interview season (**Fig. 1F**).

**Figure 1:**
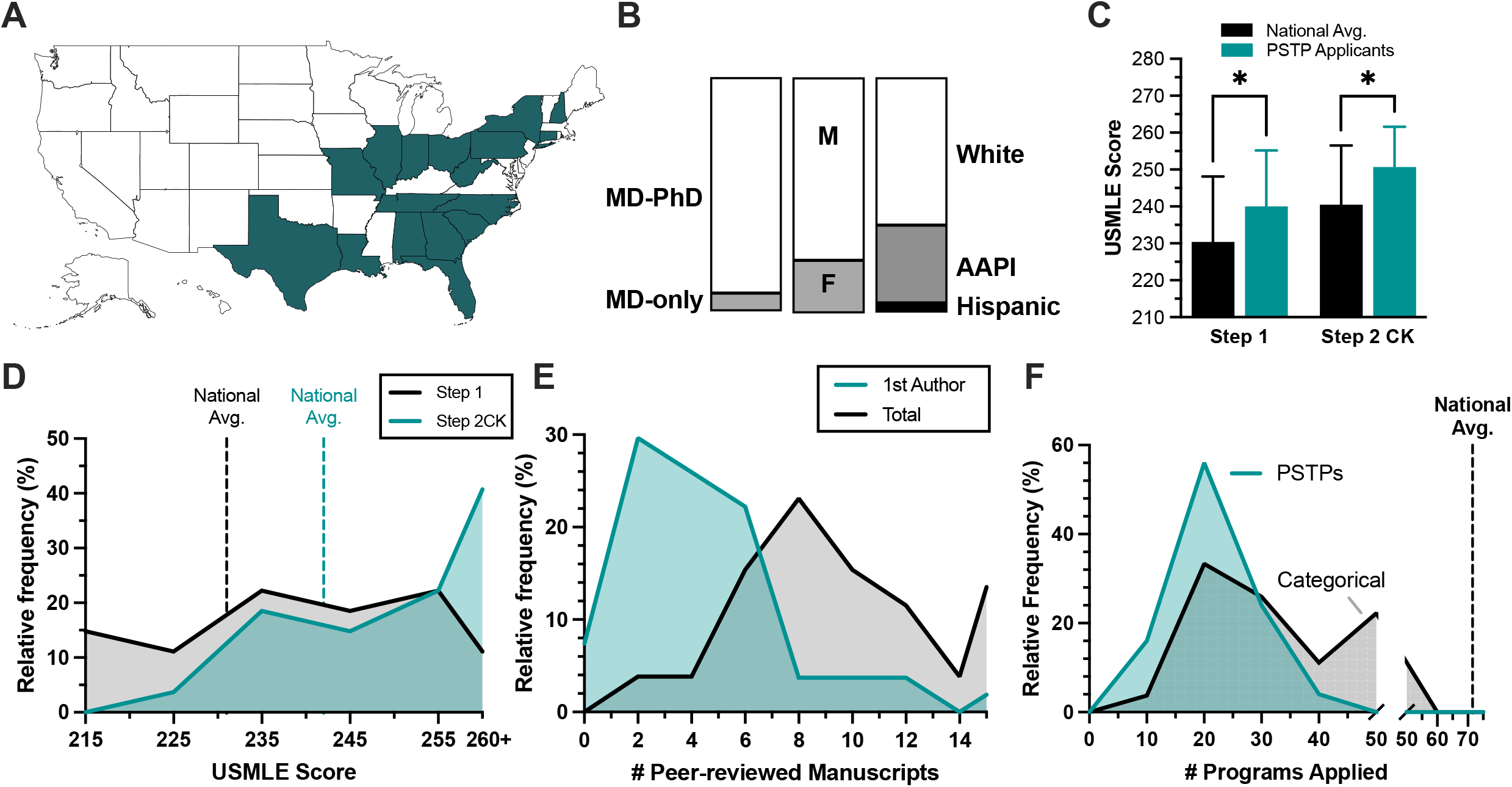
Applicant characteristics and academic performance. **(A)** geographical distribution of medical schools from which 24/27 survey respondents originated, excluding the 3 international medical graduates. **(B)** Proportional bar blot of applicant training background, gender identity, and racial identity. **(C)** Bar plot of USMLE Step 1 and Step 2 CK scores of respondents, also including the national average of graduating seniors (2022)*. **(D)** Histogram illustrating the relative frequency of USMLE Step 1 (grey) and Step 2CK (green) score distribution, with national averages reported within the AAMC 2022 post-match survey. **(E)** Histogram of peer-reviewed publications at time of ERAS submission for first-authored (green) and co-authored scientific manuscripts (grey) for PSTP applicants. **(F)** Histogram depicting the number of PSTPs (green) and categorical programs (grey) to which respondents applied. *Statistical significance at P < 0.05 using 2-way ANOVA and Šidák multiple comparisons testing, reported as mean ± SD.

### Applicants’ perceptions of desirable traits

To better understand the priorities guiding applicants’ ranking preferences, we asked respondents to weigh the value of various program-related features. Despite the broad fellowship interests represented (**Fig. 2A**), 92.6% (25/27) respondents viewed “physician-scientist community” as the most important quality of a PSTP, with 74% (20/27) also citing “structured mentoring” (**Fig. 2B**). By contrast, only half (55.6%) of respondents considered “guaranteed sub-specialty fellowship” to be an important factor influencing their decision. Notably, male (n = 22) applicants tended to value a guaranteed fellowship more highly relative to female (n = 5) applicants (*P* = 0.07). By contrast, female respondents considered access to family and/or child support (i.e. healthcare insurance, on-site childcare facilities, family, etc.) as a more important determinant of their final rank list (*P* = 0.03, **Fig. 2C**). Free response comments attributed fellowship competitiveness (e.g. cardiology), personal stability, and professional continuity when prioritizing the guaranteed fellowship.

**Figure 2:**
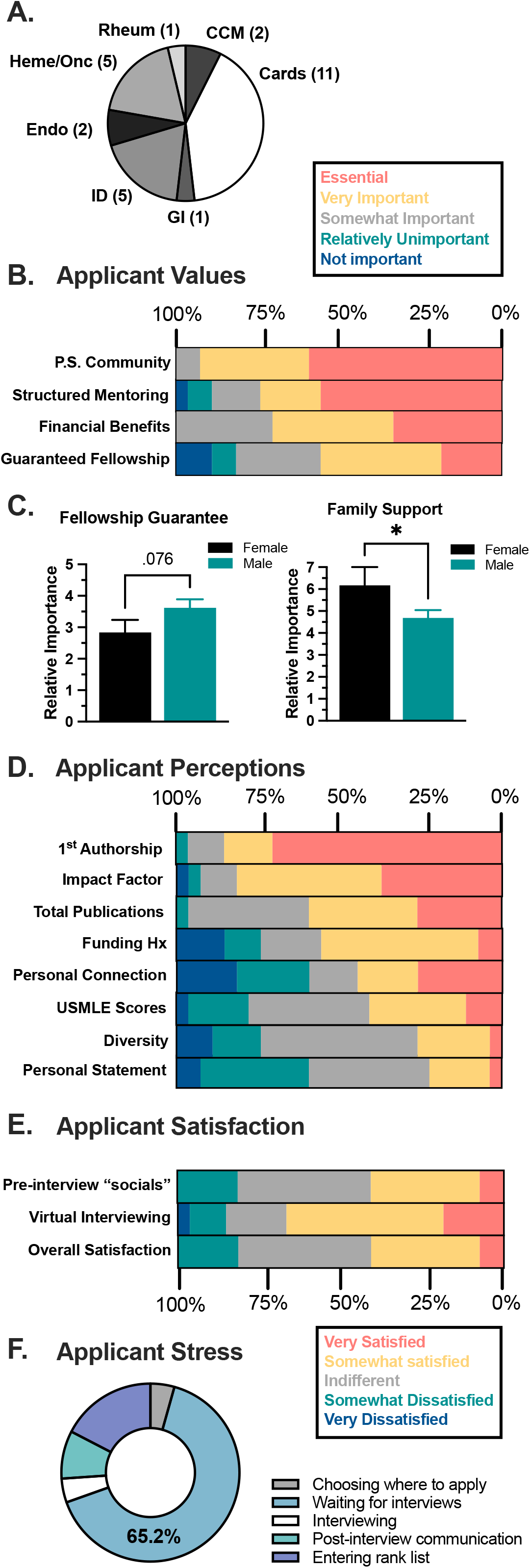
Applicant values and perceptions. **(A)** Pie chart of specialty interests of the 27 PSTP applicants who participated in the survey; the total number of respondents expressing a specialty interest is included parenthetically. **(B)** Proportional bar graphs evaluating factors of a PSTP that respondents found most important. **(C)** Gender differences in relative importance of the guaranteed fellowship and family support features offered by PSTPs when entering rank lists, differentiating female (n = 5) and male (n = 24) respondents.^#^* **(D)** respondents’ perceptions of what PSTP programs/directors most value when evaluating applicants, answered as “essential” (red), “very important” (yellow), “neutral” (grey), “relatively unimportant” (green), and “unimportant” (blue). **(E)** Proportional bar graphs of applicant satisfaction regarding virtual interviewing and overall interview experience. **(F)** Pie chart summarizing the most stressful stage of the 2022 PSTP application cycle. *Pair-wise P < 0.05. ^#^Although respondents were given non-binary gender identifiers as options, all respondents identified as either male or female.

### Application behaviors and perceptions

To better understand applicants’ perceptions of PSTP directors’ selection preferences, a series of survey items was modelled after those provided to PSTP program directors in a recent report by Gallagher *et al*.^8^ Consistent with this report, first-authorship was considered “very important” by 85% (23/27) of respondents, with 81.5% (22/27) also considering the journal impact factor in which they published as an important factor for PSTPs to consider (**Fig. 2D**). Conversely, applicants’ ERAS personal statement, USMLE scores, and applicant diversity were seen as relatively unimportant in their candidacy for PSTP admissions. Altogether, these data largely support that – apart from the thesis mentor’s letter – applicants’ perceptions of programs’ priorities aligned closely with those published by Gallagher *et al*.^8^

When asked to reflect on the interview process (**Fig. 2E**), 18/27 (66.7%) respondents favored the virtual interviewing format, though only 11/27 (40.7%) were satisfied with social aspects of the virtual interview experience. Similarly, most respondents enjoyed the financial and logistical benefits of interviewing virtually, yet most were dissatisfied with the overall interviewing experience. Free-response feedback highlighted both an inability to comprehensively assess programs via virtual encounters and a paucity of peer-peer networking opportunities.

### Predictors of match outcomes

To understand factors that may influence match outcomes, we asked survey participants about their match results. Impressively, 96.2% of respondents matched within their top-3 ranked programs, 81.5% of whom matched into a PSTP (**Fig. 3A**); by comparison, only 74.5% categorical residents matched into their top 3, with 81.5% matching into a PSTP. Among the five respondents who did not enter a PSTP, four matched at their top-ranked – and one at their #2 – categorical residency program. To begin to understand whether the qualifications described by program directors corresponded with beneficial match outcomes (i.e. matching lower on one’s rank list), we performed a correlation-based analysis of applicant metrics (**Fig. 3B**), revealing two trending factors associated with low-ranking (i.e. advantageous) matching: USMLE Step 1 (ρ = - 0.35, *P* = 0.08), and the number of programs to which a trainee applied on ERAS (ρ = 0.37, P = 0.07). Even after covariate adjustment via multiple linear regression, a significant converse association was found between the number of programs applied and the rank position to which a trainee matched (**Fig. 3C**); inverse trends were found between matched rank position and USMLE Step 1 (*P* = 0.064) and USMLE Step 2 CK (*P* = 0.072) scores (**Fig. 3D**). The number of first- and co-authorship publications did not correlate with match position (*P* = 0.34). Two applicants had no first-authored publications, both of whom matched at their #1-ranked program. Taken together, our data suggest that USMLE scores may impact match outcomes despite program messaging to the contrary, and that interviewing with more programs may not improve –perhaps even worsen – applicants’ chances of a favorable match, regardless of interview quantity. Alternatively, applicants with lower pre-interview match competitiveness may compensate by applying to more programs – accounting for the inverse correlation between rank list position and the number of applications.

**Figure 3:**
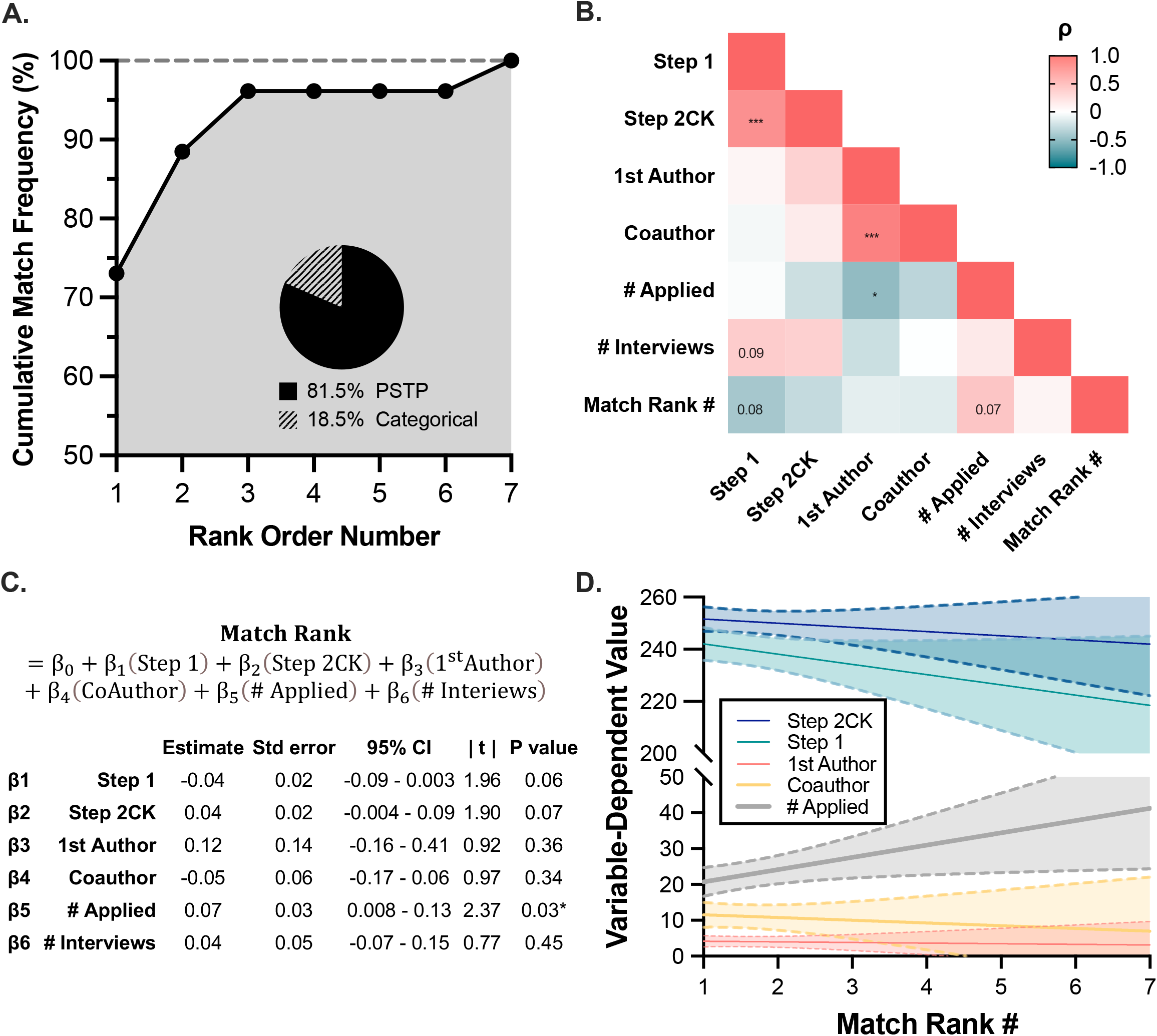
Predictors of post-match outcomes. **A**. Cumulative match frequency plotted for all survey respondents, with embedded pie graph depicting the proportion of applicants matching into a PSTP. **B**. Correlation matrix visualizing the relative association (via Pearson’s ρ) between the number of programs applied, the number of interviews received, standardized examination scores, and publication records.^#^ **C**. Table summarizing multiple linear regression modelling of the position at which respondents matched on their rank list (Match Rank #) as a function of USMLE examination scores and publication records. **D**. Plot of applicant metrics according to position on rank list matched, illustrating average ± 95% confidence interval for USMLE Step 1 and Step 2, # programs applied, # first-authorship publications, and # coauthorship publications. ^#^Significant associations via Pearson’s chi-squared test reported as *P < 0.05 and ***P < 0.001, with trending associations (P < 0.10) included.

### Post-interview communication and NRMP policy adherence

A longstanding challenge facing applicants and programs alike is navigating the interactions that follow interviews, given the bi-directional incentives to disclose ranking preferences – especially to their top choice(s) – for the purpose of influencing their match. To determine whether such factors might be influencing PSTP applicants, respondents were first inquired whether they believed a program had violated a NRMP guideline,^9^ to which nearly half (48.2%) attested to the suspicion that at least one program had infringed on NRMP policy (**Fig. 4A**), most notably by offering “misleading statements about ranking status,” “limiting post-interview communication,” and failing to “respect applicants’ right to privacy/confidentiality” (**Fig. 4B**). When asked about the most prevalent mode of post-interview communication, roughly 90% of respondents admitted that they were contacted via email by programs, with 40% meeting via virtual and/or in-person interactions (**Fig. 4C**). During these post-interview interactions, 33% admitted that they felt pressured to disclose their rank order preference, and 11% were directly asked (**Fig. 4D**). Concurrently, 63% (17/27) of respondents visited the region in which top-preferred institution was located prior to finalizing their rank list, among which 30% (8/27) met in-person with a PSTP-affiliated faculty member during their travels (**Fig. 4E**).

**Figure 4:**
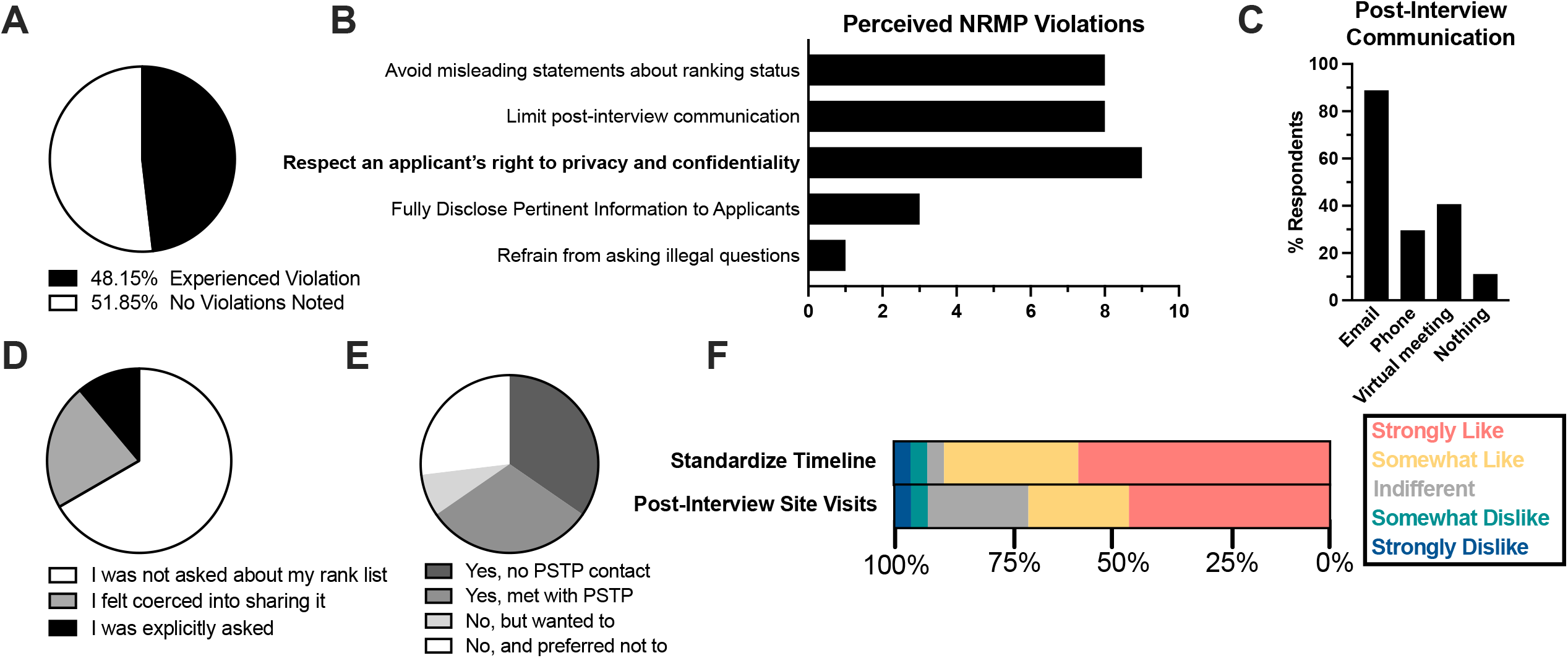
Post-interview communication and proposed application changes. **(A)** Pie chart of respondents admitting to noticing at least 1 NRMP violation by a program. **(B)** Bar plot depicting the percentage of respondents experiencing an NRMP violation. **(C)** Bar plot demonstrating the abundance of post-interview correspondence across communication modes. **(D)** Pie chart illustrating the proportion of respondents who were either asked or felt coerced into sharing their rank list with a program. **(E)** Proportion of respondents who visited a program/region, or those who wished they had visited at least one program following interviews. **(F)** Proportional bar plot of proposed changes to PSTP interview process, with answers ranging from “strongly like” to “strongly dislike.”

### Innovative strategies to overcome PSTP application

During the application and interview process, we identified several opportunities to mitigate anxiety and better inform applicants’ ranking decisions. Most popular among these was a standardized interview timeline, where nearly 85% (23/27) supported the coordination and disclosure of interview dates and invitation letters. Another 70% supported the option to meet with programs in-person via official site visits following interview season (**Fig 4F**). By contrast most applicants opposed the suggestion that programs submit final rank lists prior to site visits, citing that the benefit of influencing programs likely outweighs the potential risk. Many additional comments were submitted, offering specific examples of confusing post-interview messaging, though some respondents offered their thoughts regarding “quality of life” metrics that ultimately guided their rank list.

## DISCUSSION

Since its distribution in the early 20^th^ century, the *Flexner report* has shaped medical education into its current form by promoting standardization and evidence-based clinical training among postgraduate residencies nationwide.^10^ The mechanism of residency selection and admission has followed suit, where the Electronic Residency Application Service (ERAS) and NRMP have emerged as industry gold-standards of equity as they process over 50,000 residents annually. And yet, sub-populations of residency applicants still exist for whom “The Match” and NRMP guidelines may disfavor.^11,12^

### The “Matching” game

The Gale-Shapley algorithm, or “The Match,” is a Nobel Prize-winning project that was developed to solve the so-called “stable marriage” problem.^13^ Among its required assumptions is an acceptable degree of variability permitted by applicant pool and program size, so that the permutations needed to identify an optimal match are safeguarded by rank lists of sufficient depth for both programs and applicants. However, many physician-scientists enter residency with specialized scientific experience and career interests, where far fewer programs can offer such a suitable match. Anecdotally, those of us who opted-out of a PSTP found that no single program could offer the optimal environment at all stages of our postgraduate training. Furthermore, the 5 survey respondents that chose to enter non-PSTP categorical programs were among the most scientifically productive and academically competitive, further suggesting that PSTP selection is not a “one-size-fits-all” model, requiring more nuance than the “Match” is designed to allow.

### Cultivating community

Among the shared features of a PSTP that respondents considered most valuable was an institutional “physician-scientist community.” Yet, it is fitting to wonder whether the PSTP model universally addresses the needs and values of next-generation physician-scientists. With a median cohort size of 2 PSTP positions, to what extent can PSTPs foster community? A known contributor to long-term success is the opportunity to train in a diverse community alongside other future leaders.^14^ If unable to intrinsically sustain the needs of its trainees, programs should consider building an inter-institutional network of peers, mentors and advocates who can together offer scientific and clinical support, share perspectives, and promote trainee advancement during postgraduate residency training.

### PSTP match outcomes

Our analysis of survey-based feedback from PSTP applicants of the 2021-2022 applicant cycle challenges whether the stated priorities of PSTP selection committees represent matching outcomes. Despite the high overall match success of PSTP trainees regardless of standardized test scores, USMLE Step 1 and Step 2 CK scores were the only *positive* predictors of a favorable match. Albeit “standardized” in a numeric sense, we believe the use of USMLE scores in PSTP selection creates problems for PSTPs, misappropriating value to test-taking abilities as opposed to applicants’ potential as thought leaders of biomedical science and innovation. MD-PhD trainees must overcome the gradual inflation, or “score creep,” of national USMLE examinations,^15^ but our data suggest that this concern is on-average irrelevant for applicants applying into PSTPs. USMLE Step 1 score reporting was eliminated in 2022, a move that may disadvantage MD-PhD trainees who soon will be applying alongside MD cohorts with different USMLE score reporting.

Contrasting the “more-is-better” paradigm of pre-interview medical advising, we also found that applicants applying to more programs fared worse in match outcomes even after controlling for research productivity and USMLE board scores. This is especially noteworthy given that match outcomes were independent from the number of interviews received, suggesting that a yet-unidentified proxy of pre-interview competitiveness weighs heavily on PSTPs’ ranking decisions. Nevertheless, it may also reflect the distinct nature of PSTP selection. Most PSTP interviews span multiple days, requiring applicants to meet with categorical, fellowship, and research faculty. Preparation for interviews requires more effort as applicants must identify and submit lists of potential research mentors with whom to meet at each institution.

Consistent with our own experiences, the stress of applying into PSTPs varied widely over the course of the interview season, compounded by a multitude of questions that we as applicants inevitably faced (**Fig. 5A**). Our institutional needs for this 6–7-year program stretch beyond the clinical aspects of residency and into scientific, professional, and personal aspects of our lives. Along with the ever-changing landscape of mentors, the tailored list of suitable training sites becomes a list of one or two programs (if any) (**Fig. 3B**). The unclear expectations – combined with variable communication methods – further exacerbate this anxiety that could be mitigated through transparency and standardization.

**Figure 5:**
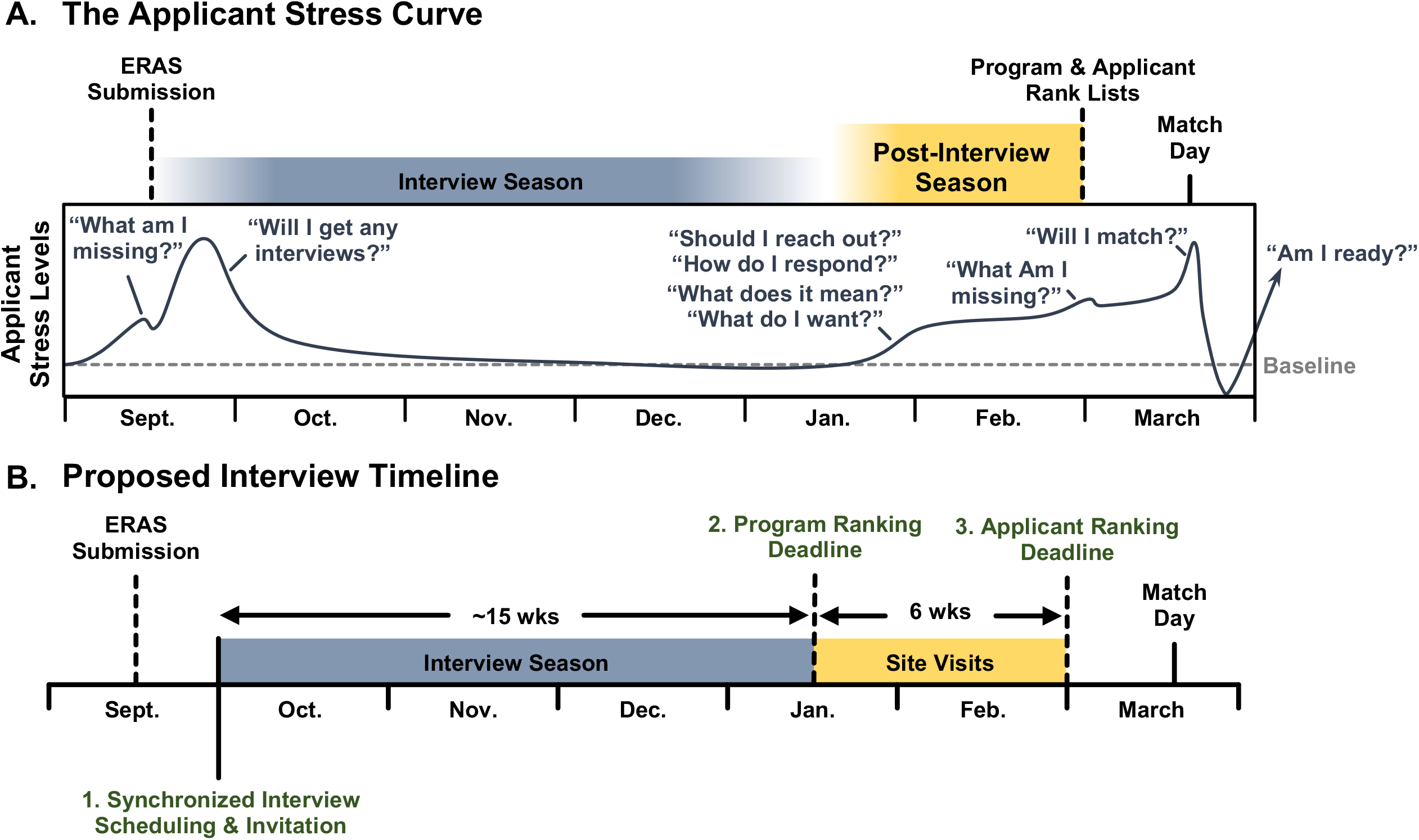
Mitigating the PSTP applicant stress curve. **(A)** Illustration of applicant stress, as assess via subjective survey-based responses. **(B)** Proposed interview timeline including synchronized interview scheduling and invitation, official site visits, and disparate program and applicant ranking deadlines.

### Post-interview communication

A noteworthy observation made from survey responses is the proportion of reciprocal NRMP violations that occur following PSTP interviews. In general, roughly 94% of categorical applicants express interest in their favorite program following interviews.^16,17^ Although only 5% of categorical programs alter their ranking in response to these “letters of interest” (LOIs),^18^ the impact of post-interview communication on PSTP ranking is probably much greater, though this has not yet been studied. Although The Match algorithm is designed to averts the need for applicants to align rank lists with those of programs, the psychological influence of post-interview “love letters” likely influences applicants’ pre-Match perceptions, as reported.^19^ The spectrum of PSTP communication styles was disorienting, with some programs requesting personal follow-up meetings, others sending PSTP acceptances, and others explicitly requesting no communication occur owing to NRMP guideline adherence. Although receiving feedback from an applicant’s preferred program mitigates match-week jitters, we feel that the unclear rules of post-interview engagement undermine its benefit.

Among the notable challenges facing applicants during interviews, the dyssynchronous application and communication timelines may be the most easily addressed. Many interview invitations arrived at unannounced times, forcing applicants to respond within minutes to schedule a non-conflicting interview date. This was especially challenging for applicants living overseas, for whom these emails arrived after working hours. For this reason, we propose the development of a centralized schedule whereby PSTPs can openly share interview dates beforehand, disclosing the dates/times at which interview invitations will be distributed.

### Study limitations

The current survey is a single-cohort study taken from the 2021-2022 application cycle, and thus may not reflect the general applicant pool on a year-to-year basis. Similarly, the small applicant pool limits the statistical power by which historically underrepresented groups could be studied to identify response patterns, shared values and/or concerns. Therefore, future studies must address the distinct concerns and/or reservations among underrepresented groups within the PSTP applicant pool.

## CONCLUSION

Our collective introduction to postgraduate physician-scientist training by way of Internal Medicine (IM) PSTP interviews underscores the importance of academic community and mentoring, which shaped our growth as both clinicians and scientists; however, applicants must navigate multiple stressors - including NRMP violations - along the way. Developing a unified PSTP interview timeline would offer a seamless application experience, whereas supplementing virtual interview encounters with non-evaluative site visits could provide equitable insights that we need to identify our next institutional home. Lastly, consistent expectations regarding post-interview communication would significantly reduce anxiety and establish ethical boundaries as applicants and programs decide how – and whom – to match. By implementing these modifications into the interview and matching process, PSTPs would further promote the entry – and retention – of physician-scientist trainees nationwide.

## Supporting information

Survey

## Data Availability

All aggregated data produced in the present study are available upon reasonable request to the authors.

## ACKNOWLEDGMENTS

We thank Dr.’s Patrick Hu (Vanderbilt University), Rebecca Baron (Brigham and Women’s Hospital), Corey G. Duke (Boston Childrens Hospital) and Randy Seay (UAB) for their insights and encouragement to publish this work.

## DATA AVAILABILITY

Aggregate data and corresponding analyses will be made available upon reasonable request to the corresponding author.

## CONTRIBUTIONS

M.E.P. designed and supervised the survey, performed the analysis, created figures, and wrote the manuscript. Y.K., B.R., M.R., and J.W. helped design the survey and edited the manuscript for clarity. M.E.P. is the guarantor of this work and thus accepts full responsibility for its integrity.

## Abbreviations

NRMP: National Residency Matching Program
PSTP: Physician Scientist Training Program
MSTP: Medical Scientist (MD-PhD) Training Program

