## Supplementary material for "Making the Match or Breaking it? Values, Perceptions, and Obstacles of Trainees Applying into Physician-Scientist Training Programs": Survey

### IM Physician-Scientist Training Program (PSTP) Applicant Survey (2021 - 2022)

In this survey, we hope to capture the applicant experience among physician-scientist trainees seeking to enter research-oriented Internal Medicine residency programs, or physician-scientist training programs (PSTPs). Thus, this survey should ONLY be completed by applicants who applied to AT LEAST 1 INTERNAL MEDICINE PHYSICIAN-SCIENTIST TRAINING PROGRAM (PSTP)** during the 2021-2022 cycle.

-----------------------------------

**Physician-scientist training programs (PSTPs) refer to residency tracks that "integrate clinical training in internal medicine RESIDENCY AND SUBSPECIALTY WITH RESEARCH ACTIVITIES over a six to seven-year period of training time." For other details, please see: https://students-residents.aamc.org/md-phd-dual-degree-training/training-physician-scientist-internal-medicine

-----------------------------------

Your participation in this research study is strictly voluntary. You may choose not to participate. If you decide to participate in this research survey, you may withdraw at any time. If you decide not to participate in this study or if you withdrawl from participating at any time, you will not be penalized.

The procedure involves filling an online survey that will take approximately 10 minutes or less to finish. Your responses will be confidential and we do not collect identifying information such as your name, email address or IP address. The survey questions will be about your application and interview experiences to PSTPs during the 2021-2022 residency interview cycle.

We will do our best to keep your information confidential. All data is stored in a password protected electronic format. To help protect your confidentiality, the surveys will not contain information that will personally identify you. The results of this study will be reported as aggregate values only, and for scholarly purposes only.

If you have any questions about the research study, please contact Mark Pepin. This research involves anonymous survey responses, and is exempt from IRB procedures for research involving human subjects.

ELECTRONIC CONSENT: Please select your choice below.

Clicking on the "agree" button below indicates that:

• you have ready the above information
• you voluntarily agree to participate
• you are at least 18 years of age





If you do not wish to participate in the research study, please decline participation by clicking on the "I disagree" button.

1. *Mark only one oval.*

   - I agree
   - I disagree

"Tell us a bit about yourself" (1/6)

2. Expected Degrees (before residency)

   *Mark only one oval.*

   - MD
   - MD, PhD
   - MD, MS
   - MD, MPH
   - MD + Other Graduate

3. In which state did you attend medical school?

   *Mark only one oval.*

   - I attended outside of the continental U.S.
   - Alabama
   - Alaska
   - Arizona
   - Arkansas
   - California

- Colorado
- Connecticut
- Delaware
- Florida
- Georgia
- Hawaii
- Idaho
- Illinois
- Indiana
- Iowa
- Kansas
- Kentucky
- Louisiana
- Maine
- Maryland
- Massachusetts
- Michigan
- Minnesota
- Mississippi
- Missouri
- Montana
- Nebraska
- Nevada
- New Hampshire
- New Jersey
- New Mexico
- New York
- North Carolina
- North Dakota
- Ohio





- Oklahoma
- Oregon
- Pennsylvania
- Rhode Island
- South Carolina
- South Dakota
- Tennessee
- Texas
- Utah
- Vermont
- Virginia
- Washington
- West Virginia
- Wisconsin
- Wyoming

4. Were any of your degrees obtained overseas?

*Mark only one oval.*

- Yes
- No

5. To which gender do you identify?

*Mark only one oval.*

- Female
- Male
- Transgender Female
- Transgender Male
- Prefer not to share
- Other: ____________

6. Are you of Hispanic, Latino, or Spanish origin?

*Mark only one oval.*

- Yes
- No
- Prefer not to share

7. How would you describe your race?

*Mark only one oval.*

- American Indian or Alaska Native
- Asian
- Black or African American
- Native Hawaiian or Other Pacific Islander
- White
- Prefer not to share
- Other: ____________





8. Are you currently living with a disability?

*Mark only one oval.*

- Yes
- No
- Prefer not to share

9. Step 1 Score Range (will not be shared)

*Mark only one oval.*

- < 210
- 210 - 220
- 220 - 230
- 230 - 240
- 240 - 250
- 250 - 260
- > 260

10. Step 2 CK Score Range (will not be shared)

*Mark only one oval.*

- < 210
- 210 - 220
- 220 - 230
- 230 - 240
- 240 - 250
- 250 - 260
- > 260

11. Academic Honor Society membership

*Check all that apply.*

- [ ] Alpha Omega Alpha (AOA) member
- [ ] Gold Humanism Honor society member
- [ ] My medical school does not have AOA and/or Gold Humanism Society

12. How many first-authored, peer-reviewed manuscripts did you have when you applied?

_______________

13. How many peer-reviewed manuscripts did you have when you applied?

_______________





14. Did you obtain extramural (e.g. NIH F-series) funding during graduate non-medical (PhD, MPH etc...) training?

*Mark only one oval.*

- Yes
- No
- N/A

15. What MOST attracted you to apply for a PSTP?

*Mark only one oval.*

- Protected research time
- Guaranteed fellowship
- Guaranteed research funding
- Physician-scientist community
- Structured mentoring
- Opportunity to "Fast-track" into postdoctoral research
- Other: _______________

16. How important was it for PSTPs to offer a GUARANTEED FELLOWSHIP position?

*Mark only one oval.*

| | 1 | 2 | 3 | 4 | 5 | |
|---|---|---|---|---|---|---|
| Not Important | | | | | | Essential |

17. ^ Why / why not?

18. How important is STRUCTURED CAREER MENTORING within PSTPs? (via PSTP-organized events, scheduling, etc...)

*Mark only one oval.*

| | 1 | 2 | 3 | 4 | 5 | |
|---|---|---|---|---|---|---|
| Not Important | ⬭ | ⬭ | ⬭ | ⬭ | ⬭ | Essential |

19. How important is it for PSTPs to offer FINANCIAL BENEFITS (research, stipend, etc...)?

*Mark only one oval.*

| | 1 | 2 | 3 | 4 | 5 | |
|---|---|---|---|---|---|---|
| Not Important | ⬭ | ⬭ | ⬭ | ⬭ | ⬭ | Essential |





20. How important is it to have a community of PHYSICIAN-SCIENTIST TRAINEES at the same institution?

*Mark only one oval.*

| | 1 | 2 | 3 | 4 | 5 | |
|---|---|---|---|---|---|---|
| Not Important | ⬭ | ⬭ | ⬭ | ⬭ | ⬭ | Essential |

21. What is your intended subspecialty?

*Mark only one oval.*

- Acute Care Medicine / Pulmonology
- Cardiology
- Endocrinology
- Geriatrics
- Gastroenterology
- Hematology/Oncology
- Infectious Diseases
- Nephrology
- Rheumatology
- Unsure
- None
- Other: ____________

#### The Application - Your Perspective (2/6)

Physician-scientist training programs (PSTPs) refer to residency tracks that "integrate clinical training in internal medicine RESIDENCY AND SUBSPECIALTY WITH RESEARCH ACTIVITIES over a six to seven-year period of training time." For other details, please see: https://students-residents.aamc.org/md-phd-dual-degree-training/training-physician-scientist-internal-medicine

In general, how important do YOU think the following factors were in obtaining a PSTP interview?

22. USMLE (Step 1/2CK) Scores

*Mark only one oval.*

| | 1 | 2 | 3 | 4 | 5 | |
|---|---|---|---|---|---|---|
| Not Important | ○ | ○ | ○ | ○ | ○ | Very Important |

23. Total number of peer-reviewed publications

*Mark only one oval.*

| | 1 | 2 | 3 | 4 | 5 | |
|---|---|---|---|---|---|---|
| Not Important | ○ | ○ | ○ | ○ | ○ | Very Important |

24. First-authored peer-reviewed Publication(s)

*Mark only one oval.*

| | 1 | 2 | 3 | 4 | 5 | |
|---|---|---|---|---|---|---|
| Not Important | ○ | ○ | ○ | ○ | ○ | Very Important |





25. Impact factor of peer-reviewed publication(s)

*Mark only one oval.*

| | 1 | 2 | 3 | 4 | 5 | |
|---|---|---|---|---|---|---|
| Not Important | ○ | ○ | ○ | ○ | ○ | Very Important |

26. History of prior extramural funding (F30, etc…)

*Mark only one oval.*

| | 1 | 2 | 3 | 4 | 5 | |
|---|---|---|---|---|---|---|
| Not Important | ○ | ○ | ○ | ○ | ○ | Very Important |

27. A personal connection with current faculty/program (self or advisor)

*Mark only one oval.*

| | 1 | 2 | 3 | 4 | 5 | |
|---|---|---|---|---|---|---|
| Not Important | ○ | ○ | ○ | ○ | ○ | Very Important |

28. Personal Statement

*Mark only one oval.*

| | 1 | 2 | 3 | 4 | 5 | |
|---|---|---|---|---|---|---|
| Not Important | ○ | ○ | ○ | ○ | ○ | Very Important |

29. Diversity

*Mark only one oval.*

| | 1 | 2 | 3 | 4 | 5 | |
|---|---|---|---|---|---|---|
| Not Important | ◯ | ◯ | ◯ | ◯ | ◯ | Very Important |

30. How much total time would you estimate spending on your application materials (in hours)?

_______________

31. To how many PSTPs did you apply?

_______________

32. To how many residency programs (PSTP or otherwise) did you apply, in total?

_______________

PSTP Interviews (3/6)

Physician-scientist training programs (PSTPs) refer to residency tracks that "integrate clinical training in internal medicine RESIDENCY AND SUBSPECIALTY WITH RESEARCH ACTIVITIES over a six to seven-year period of training time." For other details, please see: https://students-residents.aamc.org/md-phd-dual-degree-training/training-physician-scientist-internal-medicine

33. How many PSTP interview invitations did you receive?

_______________





34. How many PSTP interviews did you cancel?

_______________

35. If you canceled a PSTP interview, why?

*Check all that apply.*

- [ ] Overlapping Interview date(s)
- [ ] Already had enough interviews
- [ ] Lost interest in the program(s)
- [ ] Other: _______________

36. What is your overall opinion of VIRTUAL interviewing?

*Mark only one oval.*

| | 1 | 2 | 3 | 4 | 5 | |
|---|---|---|---|---|---|---|
| Strongly Dislike | ◯ | ◯ | ◯ | ◯ | ◯ | Strongly Like |

37. Overall, how satisfied were you with program-organized interactions with residents (pre-interview social, interview day, second-look, etc...)?

*Mark only one oval.*

| | 1 | 2 | 3 | 4 | 5 | |
|---|---|---|---|---|---|---|
| Not satisfied | ◯ | ◯ | ◯ | ◯ | ◯ | Completely satisfied |

38. Anything else you would like to say about your PSTP interview experience?

_______________________________________________
_______________________________________________
_______________________________________________
_______________________________________________
_______________________________________________

#### Post-Interview Season (4/6)

Physician-scientist training programs (PSTPs) refer to residency tracks that "integrate clinical training in internal medicine RESIDENCY AND SUBSPECIALTY WITH RESEARCH ACTIVITIES over a six to seven-year period of training time." For other details, please see: https://students-residents.aamc.org/md-phd-dual-degree-training/training-physician-scientist-internal-medicine

39. Did any PSTPs initiate communication with you after interviews?

*Check all that apply.*

- [ ] Email
- [ ] Phone call
- [ ] Virtual meeting
- [ ] Nothing
- [ ] Other: ____________________

40. Do you think post-interview communication initiated by a program influenced your ranking decision?

*Mark only one oval.*

| | 1 | 2 | 3 | 4 | 5 | |
|---|---|---|---|---|---|---|
| Definitely Not | ◯ | ◯ | ◯ | ◯ | ◯ | Definitely |





41. At any point, did you feel pressured or incentivized to share your ranking preferences with a program?

*Mark only one oval.*

- ◯ Yes, explicitly
- ◯ Yes, implied
- ◯ No

42. Which, if any, of the following NRMP codes did any PSTPs violate? If needed, please refer to https://www.nrmp.org/policy/match-code-of-conduct-programs/

*Check all that apply.*

- [ ] Ensure an interview experience that is safe, respectful, and free of harmful bias.
- [ ] Maintain ethical behavior during recruitment (e.g. no recording interviews)
- [ ] Refrain from asking illegal questions (e.g. regarding race, marital/parental status, or sexual orientation)
- [ ] Fully Disclose Pertinent Information to Applicants (i.e. criteria used for vetting applications)
- [ ] Respect an applicant's right to privacy and confidentiality (ranking preferences, other programs etc...)
- [ ] Do not require second visits
- [ ] Limit post-interview communication, and avoid misleading statements about ranking status
- [ ] Programs Rank with integrity (i.e. applicant's merit and alignment with the institution/program)

43. Did you visit with any PSTPs (or surrounding area) IN-PERSON following your interview?

*Mark only one oval.*

- Yes, and I met with someone affiliated with the PSTP
- Yes, but I did not meet with anyone affiliated with the PSTP
- No, but I wanted to visit
- No, and I preferred not to visit
- Other: ____________________

#### The Match (5/6)

For the purpose of this survey, physician-scientist training programs (PSTPs) refer to residency tracks that integrate clinical training in internal medicine RESIDENCY AND SUBSPECIALTY with research activities over a six to seven-year period of training time. For other details, please see: https://students-residents.aamc.org/md-phd-dual-degree-training/training-physician-scientist-internal-medicine

44. Where on your rank list did you match?

____________________

45. Did you match into a PSTP?

*Mark only one oval.*

- Yes
- No

46. In which state is your residency program located?

*Mark only one oval.*

- Alabama
- Alaska





- Arizona
- Arkansas
- California
- Colorado
- Connecticut
- Delaware
- Florida
- Georgia
- Hawaii
- Idaho
- Illinois
- Indiana
- Iowa
- Kansas
- Kentucky
- Louisiana
- Maine
- Maryland
- Massachusetts
- Michigan
- Minnesota
- Mississippi
- Missouri
- Montana
- Nebraska
- Nevada
- New Hampshire
- New Jersey
- New Mexico
- New York

- North Carolina
- North Dakota
- Ohio
- Oklahoma
- Oregon
- Pennsylvania
- Rhode Island
- South Carolina
- South Dakota
- Tennessee
- Texas
- Utah
- Vermont
- Virginia
- Washington
- West Virginia
- Wisconsin
- Wyoming

47. How important were research opportunities/community in your ranking preferences?

*Mark only one oval.*

| | 1 | 2 | 3 | 4 | 5 | 6 | 7 | |
|---|---|---|---|---|---|---|---|---|
| Not Important | | | | | | | | Most Important |





48. How important was the institution's reputation in your ranking preferences?

*Mark only one oval.*

| | 1 | 2 | 3 | 4 | 5 | 6 | 7 | |
|---|---|---|---|---|---|---|---|---|
| Not Important | | | | | | | | Most Important |

49. How important was the resident community when entering your ranking preferences?

*Mark only one oval.*

| | 1 | 2 | 3 | 4 | 5 | 6 | 7 | |
|---|---|---|---|---|---|---|---|---|
| Not Important | | | | | | | | Most Important |

50. How important was the program support & leadership when entering your ranking preference

*Mark only one oval.*

| | 1 | 2 | 3 | 4 | 5 | 6 | 7 | |
|---|---|---|---|---|---|---|---|---|
| Not Important | | | | | | | | Most Important |

51. When entering your ranking preferences, how important was it for a PSTP to guarantee fellowship?

*Mark only one oval.*

| | 1 | 2 | 3 | 4 | 5 | 6 | 7 | |
|---|---|---|---|---|---|---|---|---|
| Not Important | | | | | | | | Most Important |

52. How important was institutional family/child/personal support (proximity to family, health insurance, daycare, lactation room, etc...) when entering your ranking preferences?

*Mark only one oval.*

| | 1 | 2 | 3 | 4 | 5 | 6 | 7 | |
|---|---|---|---|---|---|---|---|---|
| Not Important | ◯ | ◯ | ◯ | ◯ | ◯ | ◯ | ◯ | Most Important |

53. How important was “gut feeling” when entering your ranking preferences?

*Mark only one oval.*

| | 1 | 2 | 3 | 4 | 5 | 6 | 7 | |
|---|---|---|---|---|---|---|---|---|
| Not Important | ◯ | ◯ | ◯ | ◯ | ◯ | ◯ | ◯ | Most Important |

Possible Modifications (6/6)





54. What was the most stressful aspect of your residency application?

*Mark only one oval.*

- ◯ Choosing where to apply
- ◯ Completing the application
- ◯ Waiting for interviews
- ◯ Responding to interview requests
- ◯ Interviewing
- ◯ Post-interview communication
- ◯ Entering rank list
- ◯ Waiting for match results
- ◯ Other: ____________________

55. If you could change only one thing about PSTP applications & interviewing, what would it be?

56. How would you feel if programs coordinated their interview invitations and scheduling to avoi conflicting dates (item #1 on figure)?

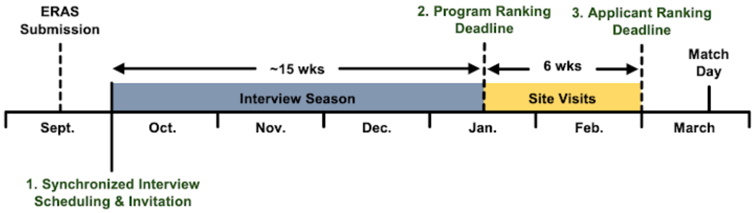


*Mark only one oval.*

| | 1 | 2 | 3 | 4 | 5 | |
|---|---|---|---|---|---|---|
| Strongly Dislike | ⬭ | ⬭ | ⬭ | ⬭ | ⬭ | Strongly Like |

57. How would you feel if PSTPs were required to submit final rank lists a few weeks BEFORE applicants? (i.e. the "site visits" below, between #2 and #3)

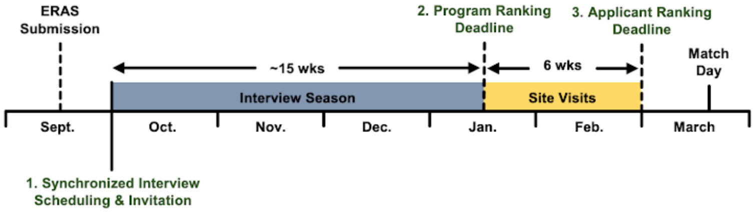


*Mark only one oval.*

| | 1 | 2 | 3 | 4 | 5 | |
|---|---|---|---|---|---|---|
| Strongly Dislike | ⬭ | ⬭ | ⬭ | ⬭ | ⬭ | Strongly Like |





58. How would you feel if PSTPs offered purely non-evaluative site visits?

*Mark only one oval.*

| | 1 | 2 | 3 | 4 | 5 | |
|---|---|---|---|---|---|---|
| Strongly Dislike | ⬭ | ⬭ | ⬭ | ⬭ | ⬭ | Strongly Like |

59. Should PSTPs participate in "The Match?"

*Mark only one oval.*

⬭ Yes
⬭ No

60. Any last thoughts/suggestions?

This content is neither created nor endorsed by Google.

Google Forms
